# Modeling layered non-pharmaceutical interventions against SARS-CoV-2 in the United States with Corvid

**DOI:** 10.1101/2020.04.08.20058487

**Authors:** Dennis L. Chao, Assaf P. Oron, Devabhaktuni Srikrishna, Michael Famulare

## Abstract

**Background:** The novel coronavirus SARS-CoV-2 has rapidly spread across the globe and is poised to cause millions of deaths worldwide. There are currently no proven pharmaceutical treatments, and vaccines are likely over a year away. At present, non-pharmaceutical interventions (NPIs) are the only effective option to reduce transmission of the virus, but it is not clear how to deploy these potentially expensive and disruptive measures. Modeling can be used to understand the potential effectiveness of NPIs for both suppression and mitigation efforts.

**Methods and Findings:** We developed Corvid, an adaptation of the agent-based influenza model called FluTE to SARS-CoV-2 transmission. To demonstrate features of the model relevant for studying the effects of NPIs, we simulated transmission of SARS-CoV-2 in a synthetic population representing a metropolitan area in the United States. Transmission in the model occurs in several settings, including at home, at work, and in schools. We simulated several combinations of NPIs that targeted transmission in these settings, such as school closures and work-from-home policies. We also simulated three strategies for testing and isolating symptomatic cases. For our demonstration parameters, we show that testing followed by home isolation of ascertained cases reduced transmission by a modest amount. We also show how further reductions may follow by isolating cases in safe facilities away from susceptible family members or by quarantining all family members to prevent transmission from likely infections that have yet to manifest.

**Conclusions:** Models that explicitly include settings where individuals interact such as the home, work, and school are useful for studying the effectiveness of NPIs, as these are more dependent on community structure than pharmaceutical interventions such as vaccination. Corvid can be used to help evaluate complex combinations of interventions, although there is no substitute for real-world observations. Our results on NPI effectiveness summarize the behavior of the model for an assumed set of parameters for demonstration purposes. Model results can be sensitive to the assumptions made about disease transmission and the natural history of the disease, both of which are not yet sufficiently characterized for SARS-CoV-2 for quantitative modeling. Models of SARS-CoV-2 transmission will need to be updated as the pathogen becomes better-understood.

## 1 Introduction

The United States has had nearly 400,000 tested or presumptive cases of COVID-19, the disease caused by the novel coronavirus SARS-CoV-2 [1]. At the time of writing, there are no approved pharmaceuticals to prevent or treat cases of SARS-CoV-2 infection. Therefore, the only available option to slow or even stop the epidemic is to use non-pharmaceutical interventions (NPIs), including school closures, workplace closures, quarantine, and other “social distancing” measures. The effectiveness of NPIs has been studied mostly in the context of seasonal and pandemic influenza, but most of the evidence is based on modeling and simulation[2–4]. However, SARS-CoV-2 is not the same as influenza. SARS-CoV-2 is more transmissible, has a longer incubation period, and may have more asymptomatic transmission. Therefore, modeling will be a useful tool for evaluating the potential effectiveness of NPIs against SARS-CoV-2 epidemics.

Here, we describe Corvid, an open-source model that simulates the spread of SARS-CoV-2 in a population. This model explicitly includes transmission in age-appropriate settings, like schools, workplaces, and homes, which allows us to study the potential impact of NPIs that target these settings. We used the model to demonstrate how these NPIs could reduce overall infections by affecting transmission in different combinations of these settings.

## 2 Results

We ran simulations in a synthetic population that represents an American metropolitan area with a population of roughly 560,000 people. In an unmitigated outbreak starting with five infected people, 81% of the population became infected when R_0_=2.6.

### 2.1 Modeling layered NPIs

To demonstrate the features of the model, we simulated the effects of using NPIs to reduce SARS-CoV-2 transmission. We first test the effectiveness of workplace-based policies then add additional “layers” of interventions that are enacted at the same time [2]. We present the impact of these interventions in figures that plot the simulated number of new symptomatic cases each day and the cumulative percentage of the population that has become symptomatic, and compare simulations with no interventions to simulations where these policies are started on either day 30 (early) or day 90 (later) of the epidemic. These simulations are meant to demonstrate features of the model and the qualitative effects of NPIs–the intervention parameters were not set to reflect any specific real-world setting.

We simulated the workplace-based policies of *liberal leave* and *work-from-home* policies. For “liberal leave”, people who normally go to work have an increased chance of staying home when feeling ill, beyond the normal tendency to stay home when ill. For “work from home”, a fraction of the employed population stays home instead of going to work. Both of these interventions can increase the amount of transmission that occurs within a family if other members of the family are at home during the day, and among people who live near their homes, but it prevents workplace transmission and the movement of the pathogen between home and work communities. In our examples, we set compliance for these policies at 50%, so that 50% of workers who would not have stayed home because of illness would stay home one day after symptom onset and 50% of all workers opt to work from home. When these policies are implemented on day 30 of the epidemic, the epidemic peak was reduced and delayed, and the final attack rate was reduced. This can be seen in the blue curves in the top panels of Figure 1, which show the simulated number of new symptomatic cases each day and the cumulative percentage of the population that became symptomatic. Enacting these policies later in the epidemic, on day 90, reduced the final attack rate but did not delay the peak, as seen in the bottom panels of Figure 1.

**Figure 1:**
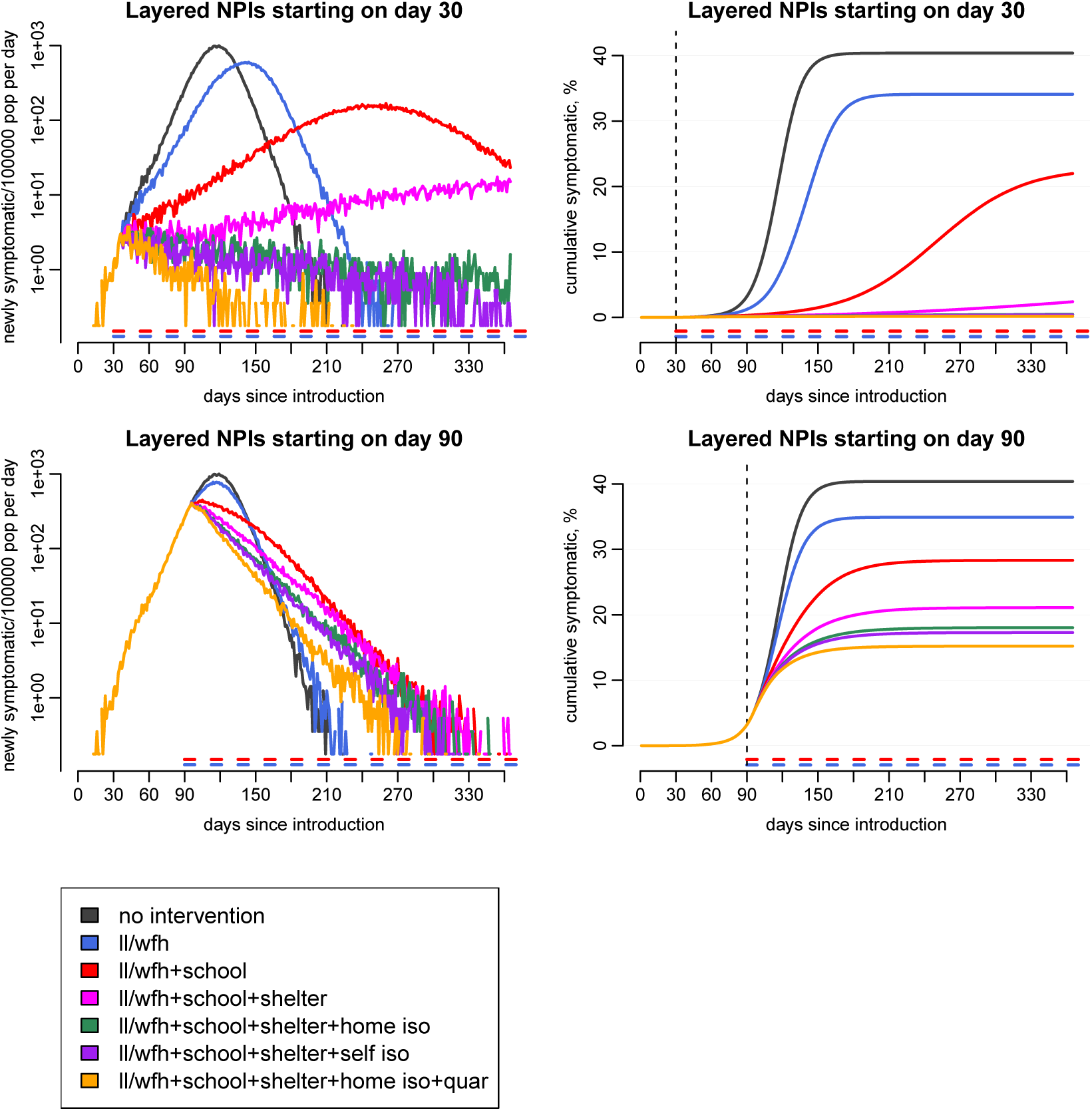
Simulating layered NPIs. NPIs were started on day 30 (top panels) or 90 (bottom panels) of the epidemic. The left plots show log-transformed numbers of newly symptomatic people per day per 100,000 population, and the right plots show cumulative % of the population that has been symptomatic. Each curve represents one stochastic simulation. The days when workplace policies are in effect is shown as a blue horizontal dashed line near the x-axis and school closures as a red line.

As expected, the workplace policies greatly reduced the number of infections occurring in workplaces and slightly increased the number occurring in the home. Figure 2 shows the number of infections that take place in the model in the home, at school, in workplaces, and outside these settings for the scenarios described above.

**Figure 2:**
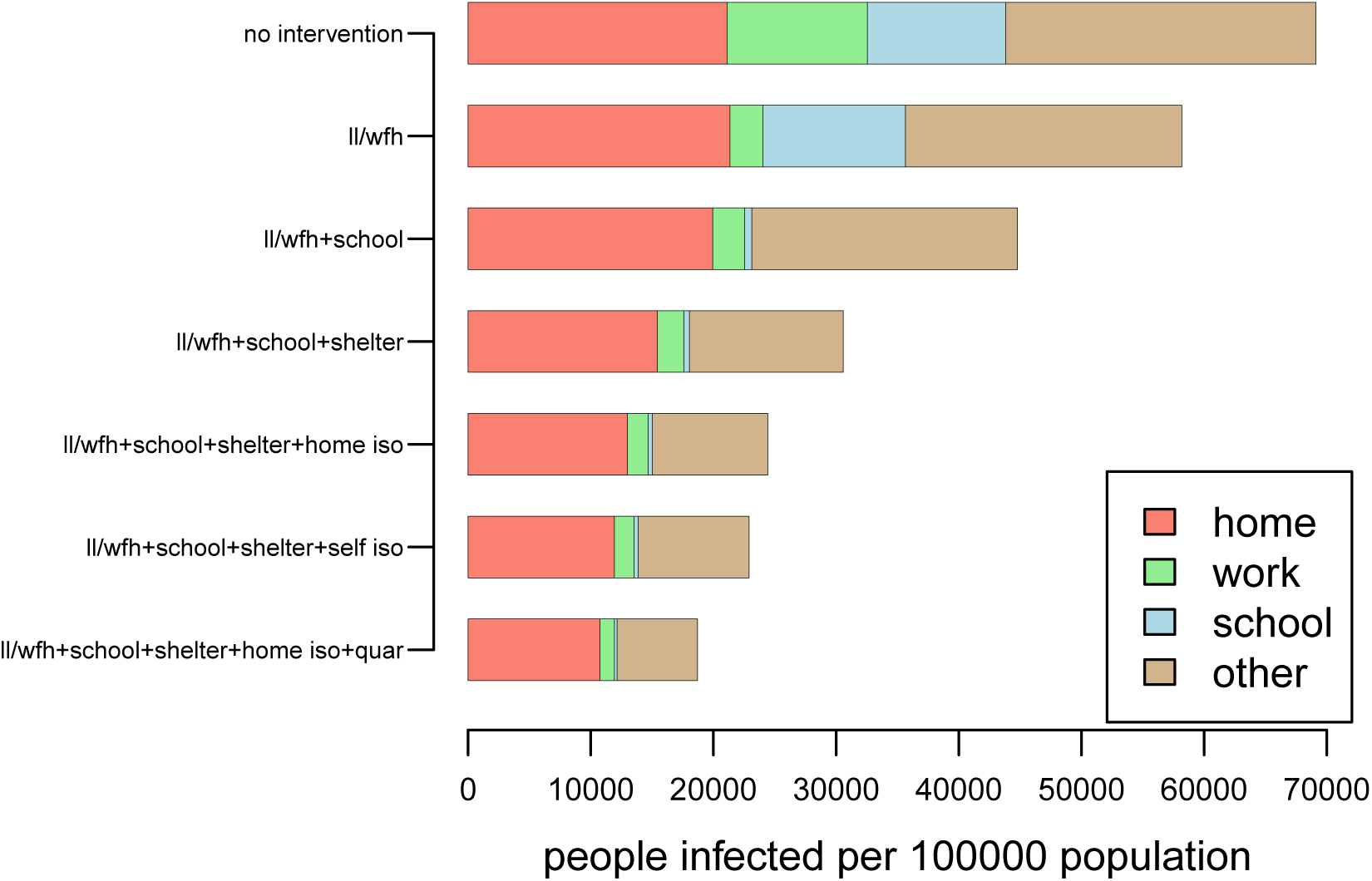
Settings of infections under layered NPIs. The bars show the number of symptomatic cases per 100,000 population infected in one of four settings: “home”, “work”, “school”, and “other”. Each row represents the number of infections that occur after day 90 during for one scenario starting on day 90 of the epidemic, corresponding to the scenarios in the bottom panels of Figure 1. Each row represents the results from one stochastic simulation.

We simulated adding school closures to these workplace policies. When schools close in the model, transmission stops occurring in schools but children have slightly more potentially infectious contacts in their communities. Preschool playgroups are not subject to school closures but transmission that occurs in them are counted towards “schools” in the model, so there will be some apparent school-based transmission after school closures. Adding school closures reduced the final attack rates even further (red curves in Figure 1). We also ran simulations in which schools re-opened while workplace-based NPIs were left in place. We observed the epidemic rebound after schools re-opened because we did not simulate scenarios that resulted in elimination (Figure 3).

**Figure 3:**
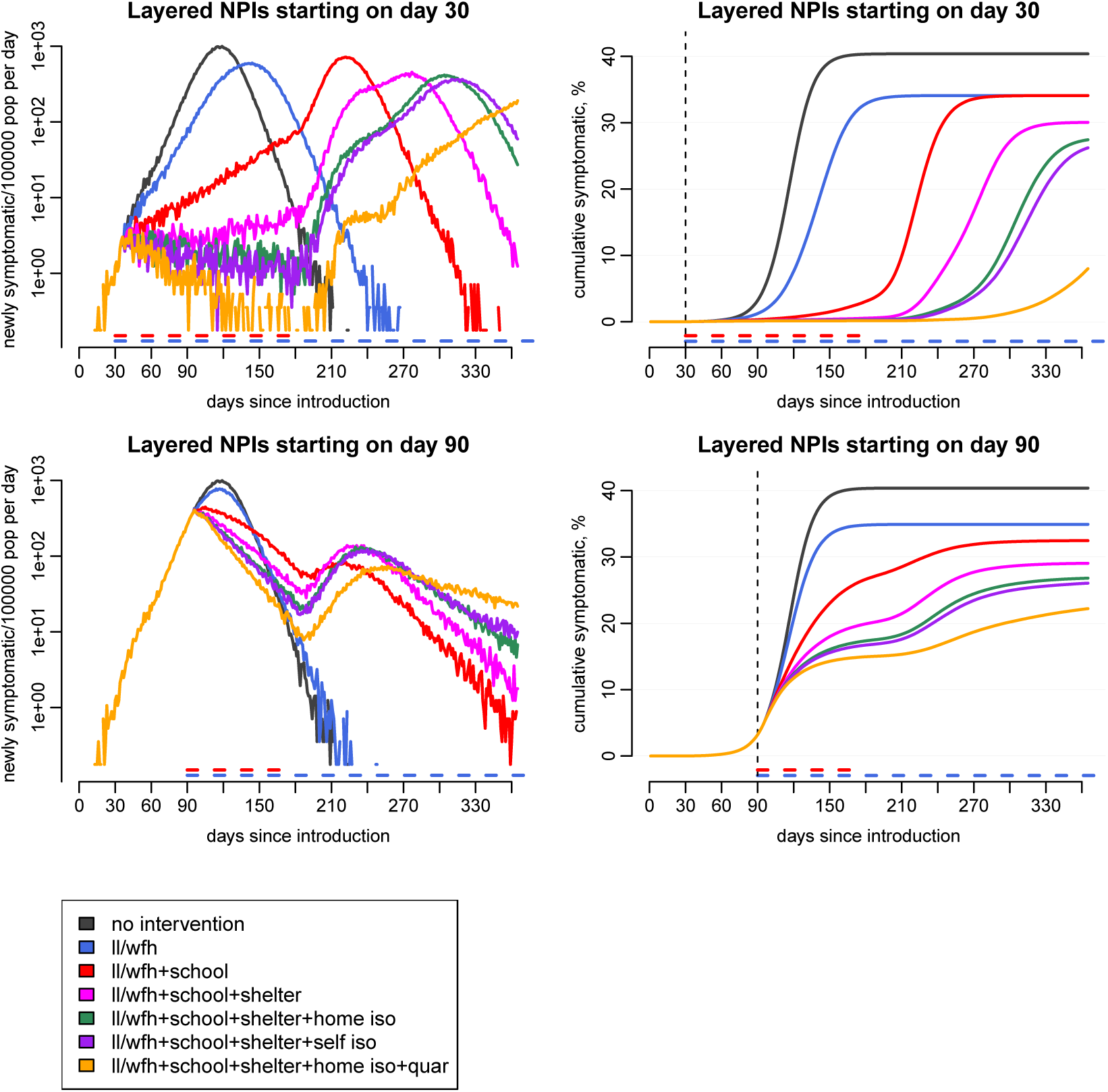
Simulating re-opening schools after school closure and other sustained NPIs. NPIs were started on day 30 (top panels) or 90 (bottom panels) of the epidemic, and schools were re-opened on day 180. The left plots show log-transformed numbers of newly symptomatic people per day per 100,000 population, and the right plots show cumulative % of the population symptomatic. Each curve represents one stochastic simulation. The days when workplace policies are in effect is shown as a blue horizontal dashed line near the x-axis and school closures as a red line.

Adding *shelter-in-place* reduced transmission even further. We modeled shelter-in-place by reducing everyone’s community contacts (contacts outside of home, work, and schools) by 30%. With schools and workplaces already closed, the shelter-in-place reduced the largest remaining component of transmission, the community (Figure 2). However, for these demonstration parameters there was still substantial transmission in the home (Figure 2 and Table 2)

**Table 2:**
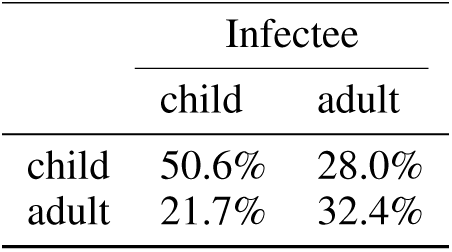
Simulated household secondary infection attack rates. The columns are the probabilities of becoming infected by age of the susceptible household member. The rows are the probabilities of an index case infecting other household members based on the age of the index case.

|  | Infectee |  |
| --- | --- | --- |
|  | child | adult |
| child | 50.6% | 28.0% |
| adult | 21.7% | 32.4% |

Finally, we added three different test-and-isolate strategies to the combination of workplace, school, and community NPIs described above. We simulated *home isolation, self isolation*, and *home isolation with household quarantine* of ascertained cases. First, we studied a best-case testing scenario in which symptomatic people are tested and results are ready immediately upon becoming symptomatic. To simulate home isolation, we assumed a person stops going to work or school after testing positive, though there is still some contact with the community during isolation. In this scenario, adding home isolation to the other NPIs can reduce transmission in the community (Figures 1– 3 and Table 1). The impact was not substantial when the other NPIs were already controlling the epidemic (bottom panels of Figure 1), but its contribution made a qualitative difference when the combined impact of all the NPIs changed the epidemic’s trajectory from a slow rise to a slow decline, leading to near-elimination (top panels of Figure 1). We simulated self isolation by blocking transmission to all contacts including household members. For this scenario where individuals are completely isolated from the susceptible population, self isolation was a little more effective than home isolation, indicating the role of secondary transmission in the household from the ascertained case. The household quarantine scenario–home isolation paired with a 14-day quarantine of all household members–was the most effective of the three strategies. We believe that quarantine of household members addresses both secondary transmission from the index case as well as the possibility of transmission from other household members that were already infected before the first symptomatic household member was detected. For all scenarios, when we included a delay in testing, introducing a 5-day delay between becoming symptomatic and beginning isolation, the impact of these isolation strategies was reduced (Figure 4). In particular, home and self isolation became equally ineffective, since cases were identified too late to prevent substantial transmission.

**Table 1:** Simulated proportion of infections by setting.

| setting | $R_0=2.6$ | $R_0=2.0$ |
| --- | --- | --- |
| school | 17.9% | 20.0% |
| work | 16.3% | 15.3% |
| home | 30.1% | 31.1% |
| community/neighbors | 35.7% | 33.6% |

**Figure 4:**
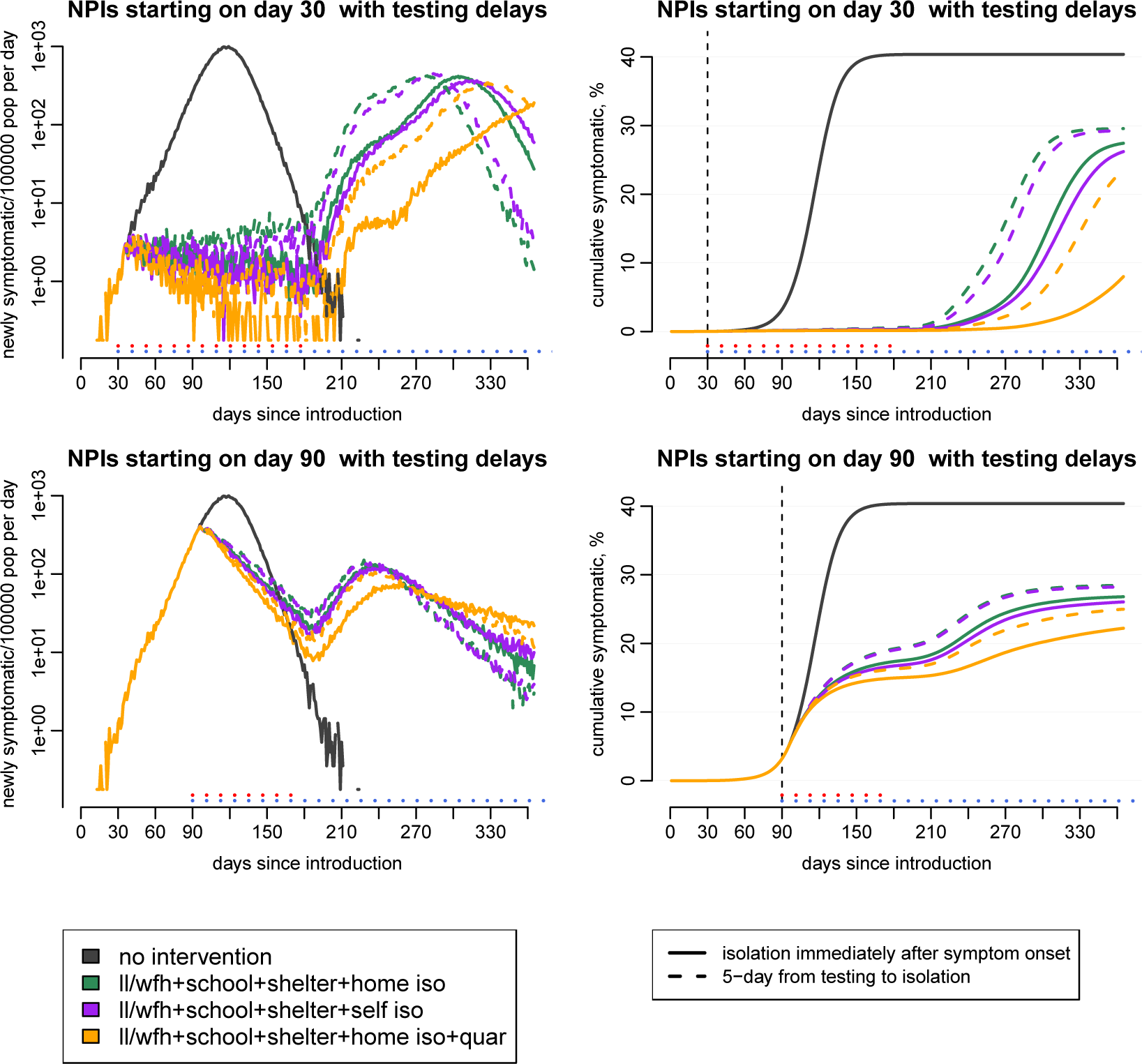
The effect of testing delays on test-and-isolate strategies. We simulated test-and-isolate strategies in which ascertainment of cases took place immediately upon symptom onset (solid lines) or 5 days after symptom onset (dashed lines). The simulations with no testing delay (solid lines) are identical to those shown in Figure 1: all NPIs start on day 30 (top panels) or 90 (bottom panels) of the epidemic, and schools are re-opened on day 180. Three different isolation strategies were modeled: home isolation, self isolation, and home isolation with family quarantine. 50% of symptomatic people were tested. The left plots show log-transformed numbers of newly symptomatic people per day per 100,000 population, and the right plots show cumulative % of the population symptomatic. Each curve represents one stochastic realization of the model. The days when workplace policies are in effect are shown as blue horizontal dotted lines near the x-axis and school closures as red lines.

### 2.2 Stochastic persistence

When many NPIs are implemented simultaneously, R_effective_ can be brought below 1.0 resulting in declining prevalence. One such example was shown in the top panels of Figure 3, in which liberal leave, work-from-home, school closure, shelter-in-place, and home isolation of 50% of ascertained cases started on day 30 and schools re-opened on day 180. Because elimination did not occur, the epidemic rebounded when schools re-opened. To demonstrate stochastic effects in the model, we ran the same scenario with different random number seeds (Figure 5). Using different random number seeds produced different numbers of cases when all interventions were in place from days 30 to 180, and these differences were amplified after schools re-opened.

**Figure 5:**
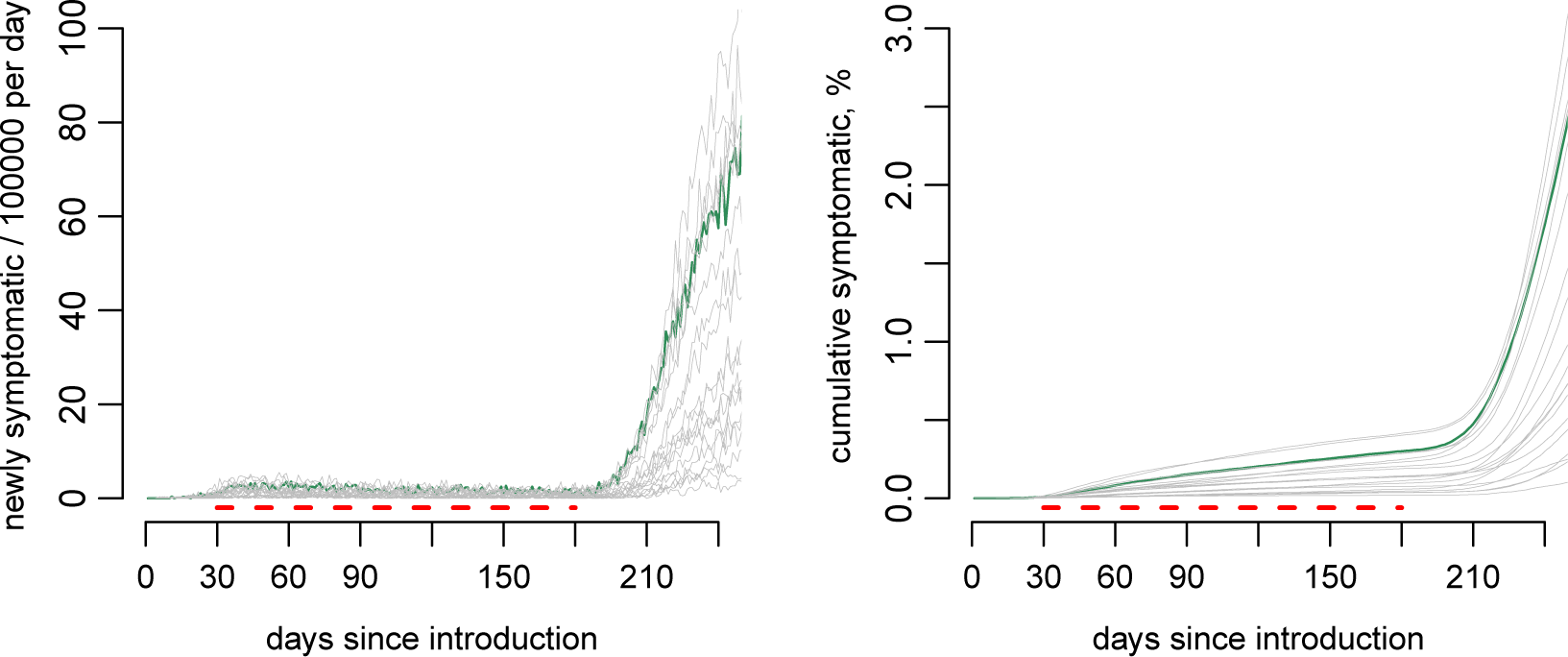
Stochastic persistence of SARS-CoV-2 until schools re-open. Liberal leave, work-from-home, school closure, community contact reduction, and home isolation of 50% of ascertained cases was started on day 30 of the epidemic, and schools were re-opened after 150 days. Each curve is one of twenty stochastic simulations with the same interventions simulated. The green lines replicate the stochastic run plotted in Figures 3 and 6. The dashed red line on the bottom indicates the school closure dates.

To understand what persistence looked like during those 150 days before schools re-opened, we analyzed transmission in one stochastic simulation run. When the interventions started on day 30, 261 people were infectious, 55 of whom were symptomatic on that day. The number of people who were infectious continued to rise for about 3 weeks (the infectious period in the model), peaking at 598 before declining. When schools re-opened on day 180, there were 315 infectious people who could re-start the epidemic when the schools opened. When school, workplace, community-targeted NPIs and home isolation were in effect, R_effective_ was 0.96. Before schools and many workplaces closed, there were chains of transmission within schools and workplaces (Figure 6, blue and green nodes). When all NPIs were in effect, a larger proportion of transmission occurred within families (Figure 6, red nodes) and between families by contacts in the community (Figure 6, brown nodes). This scenario did not include interventions that reduced transmission within households, so they may sustain the outbreak by amplifying cases that infect new households.

**Figure 6:**
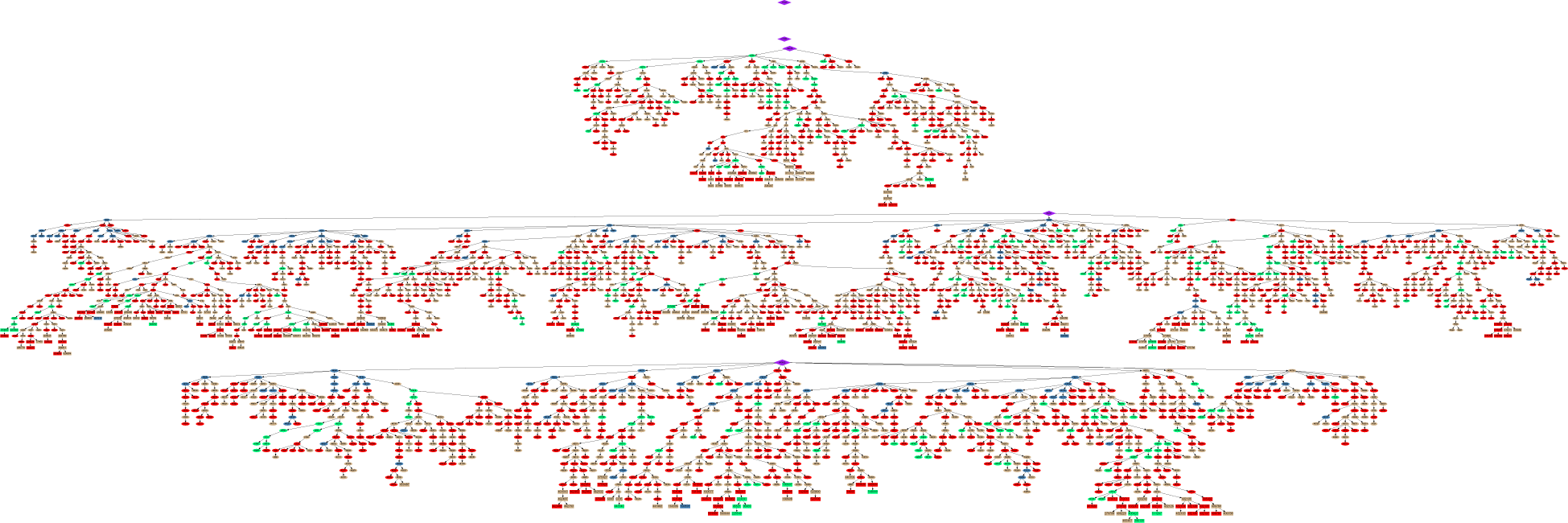
Transmission trees of an outbreak while several NPIs are in place. Liberal leave, work-from-home, school closure, shelter-in-place, and home isolation of 50% of ascertained cases were started on day 30 and schools re-opened on day 180. Ovals, rectangles, and diamonds represent individuals who were infected before the shutdown ended, and arrows indicate who infected whom but line lengths are not proportional to time between infections. Purple diamonds are the five initial cases seeded on day 0, and these are the roots of the five transmission trees that comprise this outbreak. The colors of the ovals indicate where people were infected: red for those infected by household members, blue in schools, green at work, and tan in the general community. The larger rectangles are people who were still infectious when the schools re-opened. Transmission events after schools re-opened are not shown. The small printed numbers are unique IDs for each person in the synthetic population.

When schools are closed and workplace and other contacts are reduced, there may be high transmission within households and low transmission everywhere else. The persistence of SARS-CoV-2 could be understood as transmission between households rather than transmission between individuals [5–8]. We can define a “household” analog of R_0_ to be the average number of households that a typical household infects in a totally susceptible population, and R_effective_for households to be this measure at a point in time when the population is not totally susceptible and an intervention may be taking place. When liberal leave, work-from-home, school closure, shelter-in-place, and home isolation were in effect at the levels described above, households infected 0.89 other households.

## 3 Discussion

We introduced and demonstrated the use of a new agent-based model, Corvid, to simulate a variety of NPIs against SARS-CoV-2. Simulations of single interventions help us understand their contribution to reducing transmission, but we do not consider them realistic, since interventions can affect the lives of non-targeted people. For example, if schools are closed, parents may need to stay home to care for their children, creating a *de facto* work-from-home policy. Multiple simultaneous NPIs could suppress transmission for long periods of time, but stochasticity (“bad luck”) can allow SARS-CoV-2 to persist for months under these conditions even if R_effective_ is slightly below 1. Therefore, when NPIs were relaxed, the epidemic could rebound. Although we did not include importation of cases in our simulations, the same sort of rebound could be expected if new cases arrived in the population after NPIs are relaxed. Combinations of interventions that bring R_effective_ far below 1 might be required for elimination in a shorter time frame [9]. But even if elimination does not occur, combinations of NPIs can drastically reduce the prevalence of infections, essentially “resetting” the epidemic to an earlier and more manageable state. Of course, these interventions are costly to society, so adaptively modulating them to maintain incidence within a tolerable range is an appealing option if surveillance is sufficiently broad and timely [9, 10].

Our model results indicate that a large proportion of infections could occur in the household and that the household secondary attack rates may be high. This conclusion comes from calibrating the model using published observations of influenza transmission [11–13] and extrapolating using SARS-CoV-2 characteristics. We are not yet aware of a quantitative synthesis of the evidence for household attack rates and their heterogeneity. In particular, the model currently assumes that children are a major factor in the spread of SARS-CoV-2, as they are for influenza, which has not been proven or disproven at this time [14, 15]. If children are less susceptible to infection or are less effective transmitters than adults with similar clinical presentations, it would impact the effectiveness of school closures and the rebound following re-opening.

Testing and home isolation of positive cases could serve the broader community more than the families of these cases. However, our model highlights that untested family members may often be infected before index cases are tested, so automatic home quarantine for all family members might be warranted if testing them is not possible. Because most infections in our model come from pre-symptomatic people or from people who never show symptoms, it may be impossible to prevent most infections with test-and-isolation policies unless non-symptomatic people are also isolated (e.g., quarantine of families or testing non-symptomatic people). The testing and isolation simulation results are particularly sensitive to assumptions about asymptomatic and pre-symptomatic transmission [16], neither of which is yet well-characterized for SARS-CoV-2. Another modeling study found testing with isolation to be more effective, but their model assumed less asymptomatic transmission than Corvid does currently [17]. Contact tracing might be required to control SARS-CoV-2 outbreaks [18], but could be more labor-intensive than implementing quarantine of families of ascertained cases. The proportion of SARS-CoV-2 infections that become symptomatic appears to be age-dependent [19–21], but this is both hard to measure and hard to incorporate in a model. In Corvid, the presence of symptoms drives testing of cases, and mildly symptomatic people might not be suspected of having COVID-19. We therefore need to carefully consider how to choose an appropriate symptomatic fraction to model the human behaviors that drive both disease spread and detection. More extensive testing of asymptomatic contacts would help us understand the prevalence of asymptomatic infections and the spectrum of disease more generally.

Our model has many obvious limitations. It was developed as the epidemiology of SARS-CoV-2 was becoming understood early in the pandemic. We assumed that SARS-CoV-2 could be modeled as a more transmissible influenza with a longer serial interval. The seasonality of SARS-CoV-2 transmission is not yet known and is not part of the model. Our model does not estimate hospitalizations or deaths due to COVID-19. Morbidity and mortality rate estimates are rapidly evolving and depend on local health facilities, and they can be applied to our model output of SARS-CoV-2 infections. Hospitals, healthcare workers, and other high-risk professionals are not included in our model. Finally, our work is based on an older influenza model [13], so it uses US Census data from the year 2000. The 2000 Census included a detailed commuter survey that Corvid requires to simulate movement between census tracts, so it would be difficult to update it with newer Census data.

So far, few models of SARS-CoV-2 explicitly include key settings of transmission [10, 22]. We believe that these kinds of models are required to mechanistically simulate combinations of NPIs, which would be difficult to simulate without considering how these interventions target transmission in specific settings but may increase transmission elsewhere. However, human behavior in response to NPIs is difficult to model, and we believe that model results should be interpreted cautiously. Many of the population structure assumptions in Corvid were calibrated to match expert opinion on the relative importance of a few major settings (work, school, home, neighborhood) for seasonal influenza. As governments enact social distancing measures of unprecedented scale and duration and as public awareness of the dangers of COVID-19 increase, these assumptions might not hold and models should adapt. Despite these caveats, modeling may be one of the best available guides to think through the potential effectiveness of complex combinations of NPIs.

## 4 Methods

### 4.1 Model description

Corvid is based on a previously published influenza transmission model called FluTE, described in detail in [13]. The synthetic populations and the behavior of individuals in the population are taken directly from FluTE. Disease transmission and natural history parameters were changed to match what is currently known about SARS-CoV-2. Interventions in the model were modified or added to reflect current discussions about COVID-19. The C++ source code for Corvid and the scripts used to generate the figures in this manuscript are available at https://github.com/dlchao/corvid. Version 0.5 of Corvid was used to produce the results in this manuscript. Analyses were performed and plots were made using the R programming language version 3.6.1 [23] and GraphViz version 2.40.1 (https://www.graphviz.org/).

In brief, Corvid creates communities of about 2000 people, and to simulate larger populations, many little communities are created and connected through commuting patterns. Synthetic populations for US locations are created by generating enough communities to represent each census tract, with a population size matching the 2000 US Census data, and linking those communities using commuter data from that Census. Unfortunately, more recent censuses did not offer the commuting data. Individuals are in 5 age bins: pre-school, school-aged, young adult, older adult, and elderly. Individuals are generated by populating the communities with families with the household sizes and age distribution drawn from the 1% public use microdata sample (PUMS) of the 2000 Census (https://www.census.gov/main/www/pums.html). The use of PUMS data was introduced to FluTE after the [13] publication.

The model runs in discrete half-day time steps, representing day and night. During the day, people go to institutions (mixing groups) appropriate to their age, and at night they return to their families (where ages mix). Working-age adults may commute to different census tracts during the day. We do not distinguish between weekdays and weekends, so people go to school and work every day. Susceptible people become infected when they are in the same setting as infected people. Susceptibles also have a small chance of infection if they are in the same community at the same time as an infectious person. Children and adults are equally susceptible to infection, but children tend to have stronger “contacts” in the model so are more likely to infect and become infected than adults. Upon infection, a person may become infectious starting the next day (regardless of symptom status) and their level of infectiousness can change daily (e.g., exponential, log-normal, etc). A fraction of infectious people become symptomatic after an incubation period, which is specified as a CDF for flexible parameterization (e.g., can be fixed incubation time, normal, Weibull, etc). Symptomatic people may choose to stop going to work or school because of illness. Public health interventions, such as home quarantine or school closures, may be based on the cumulative prevalence of detected symptomatic cases. Social distancing is implemented by closing settings, like schools, to stop transmission but this is partially offset by these individuals going elsewhere. Much more detail about model structure and assumptions is in [13].

We ran Corvid in a synthetic population based on 2000 US Census data for the Seattle metropolitan area, which we take to represent a typical large city in the US. The synthetic population has 563,484 people residing in 124 census tracts (Figure 7). Tracts have 1181 to 9003 people living in them, and 150 to 36,823 people working in them.

**Figure 7:**
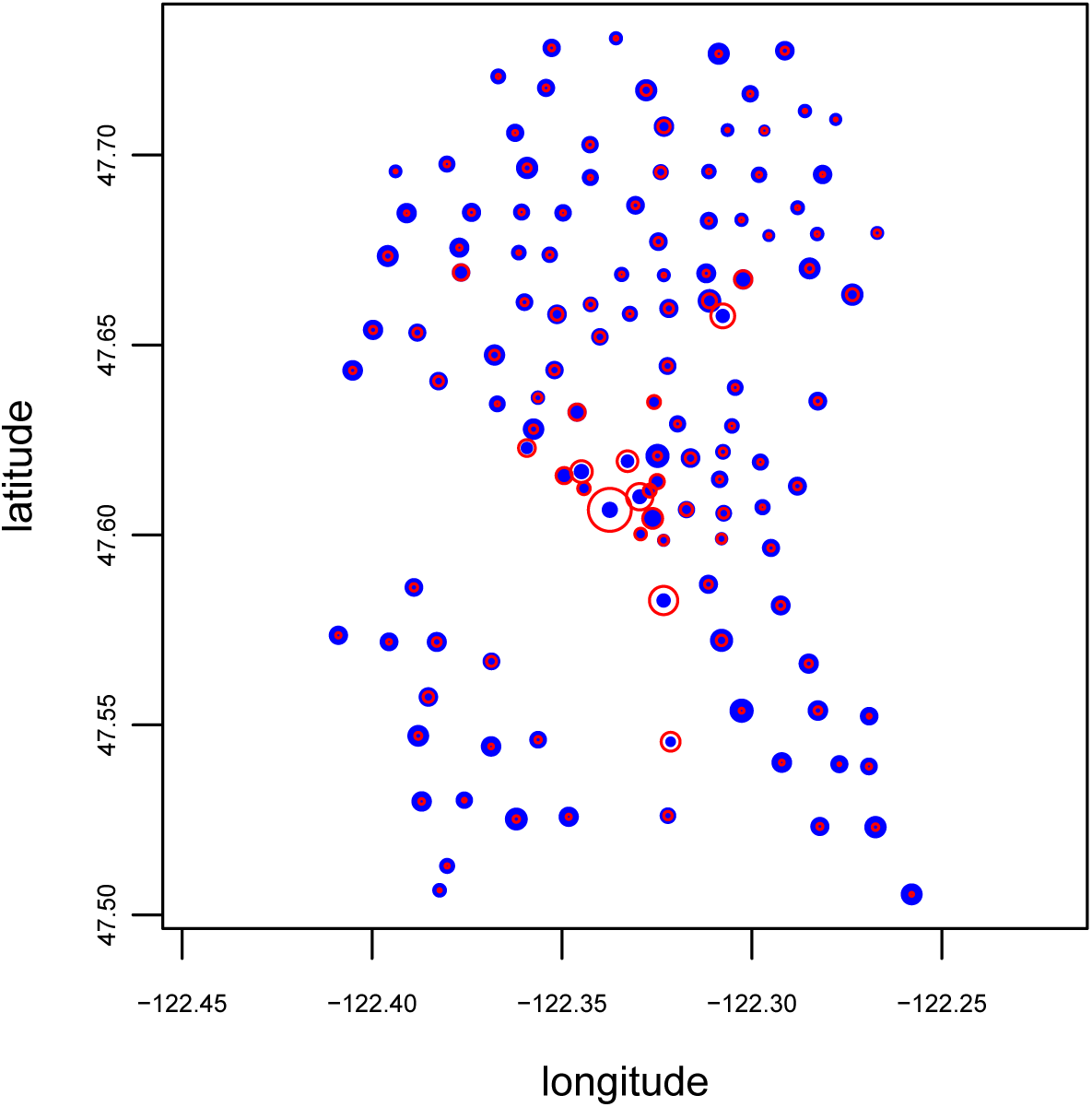
Map of the census tracts for the synthetic Seattle population. Dot size proportional to resident population. Red circle size is proportional to the number of people who work in each tract. Tract population sizes based on Census data from the year 2000.

### 4.2 Model calibration for SARS-CoV-2 transmission

Corvid was calibrated to simulate SARS-CoV-2 transmission. Where possible, parameters were derived from the literature. Other disease parameters were taken from influenza (FluTE’s default parameters). Corvid users should update and re-calibrate as more information about SARS-CoV-2 becomes available.

We assume that half of people infected will become symptomatic. This is a provisional assumption until we can obtain more reliable estimates.

For the incubation period distribution (duration from infection to showing symptoms), we used the log-normal fit from [24] who used data from Shenzhen. They found parameters of log(mean)=1.621 and log(sd)=0.418 (Figure 8). There are reports that viral detection peaks a few days after symptoms. [25] found pharyngeal shedding to peak 4 days after symptoms using throat swabs. Therefore, we used the incubation period distribution and added 4 to the mean (the log(mean) of the distribution is log(exp(1.621)+4)). That makes shedding peak 4 days after the median incubation period (Figure 8). It might make more sense for shedding to peak right after symptoms appear in an individual instead of independent of symptom onset. We assume that shedding lasts for 21 days, though the level of shedding is low by the end of the infectious period. We make asymptomatic individuals half as infectious as symptomatic. So if a person never becomes symptomatic, infectiousness is simply half that of a symptomatic individual over time. If a person becomes symptomatic five days after becoming infected, then for the first five days that person is less infectious and after that approximately twice as infectious. To our knowledge, there is no direct evidence that asymptomatic people are less infectious than symptomatic, but we assume that the symptoms like sneezing mechanically spread disease. There are some hints that asymptomatic have viral loads similar to symptomatic [26].

**Figure 8:**
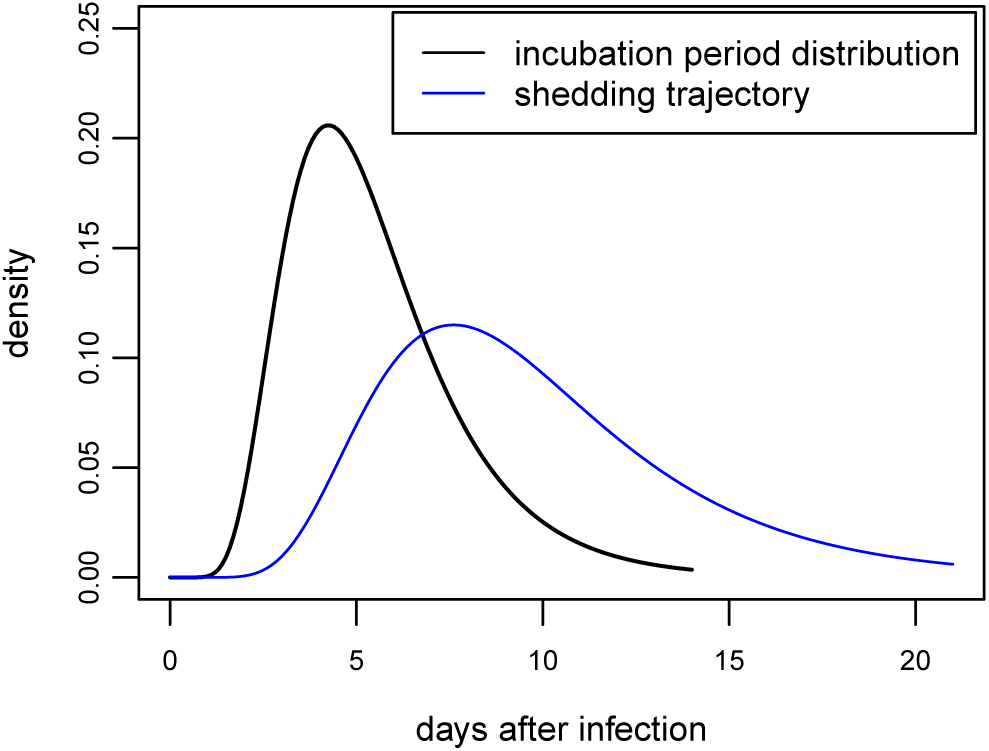
Incubation period distribution and shedding trajectory in the Corvid model. Black curve is the log-normal distribution of incubation periods from [24], truncated at day 14. Incubation periods are drawn from this distribution in Corvid. The blue curve is shedding over time in Corvid, which is a log-normal distribution with a higher mean but same standard deviation as the incubation distribution. The distribution truncated at day 21. In Corvid, people start shedding the day after infection regardless of symptomatic status.

We calibrate R_0_ by counting the number of people an index case can infect in the model. This captures not only shedding kinetics, but behavior in response to illness (tendency to stay home) and age-dependent mixing patterns. We set transmissibility of coronavirus in the model by multiplying contact probabilities by the scalar *β*, which we can set in the model. We derive the relationship between R_0_ and *β* by running the model with different values of *β* and counting the number of people an index case infects in a fully susceptible population. For each value of *β* that we test, we infect one randomly selected individual in the population. We fit a line through the average number of secondary cases for each value of *β* tested.

The number of people infected depends on the age of the index case. For example, if an index case is a school-aged child (indicated by “1” on the plot), the number of secondary cases is higher than for other age groups. Also, the secondary cases are not representative of the general population – school-aged children are also over-represented among infectees. R_0_ should be defined as the number of secondary cases generated by a “typical” case. Therefore, we tally the number of secondary cases generated by all index cases across runs to get a better idea of who is more likely to be a “typical” infecter. We weight the relationship based on the proportion of secondary cases in each age bin; we put more weight on the high transmission from school children than the lower transmission from adults in our R_0_ calculation (Figure 9). For the default model, we set R_0_ to be 2.6 (*β*=0.168206), but we also test R_0_=2.0. These estimates are in line with the literature [26–28]. We often initialize the model with more than one infected individual. When you seed with one, there is a high probability that an epidemic does not take off.

**Figure 9:**
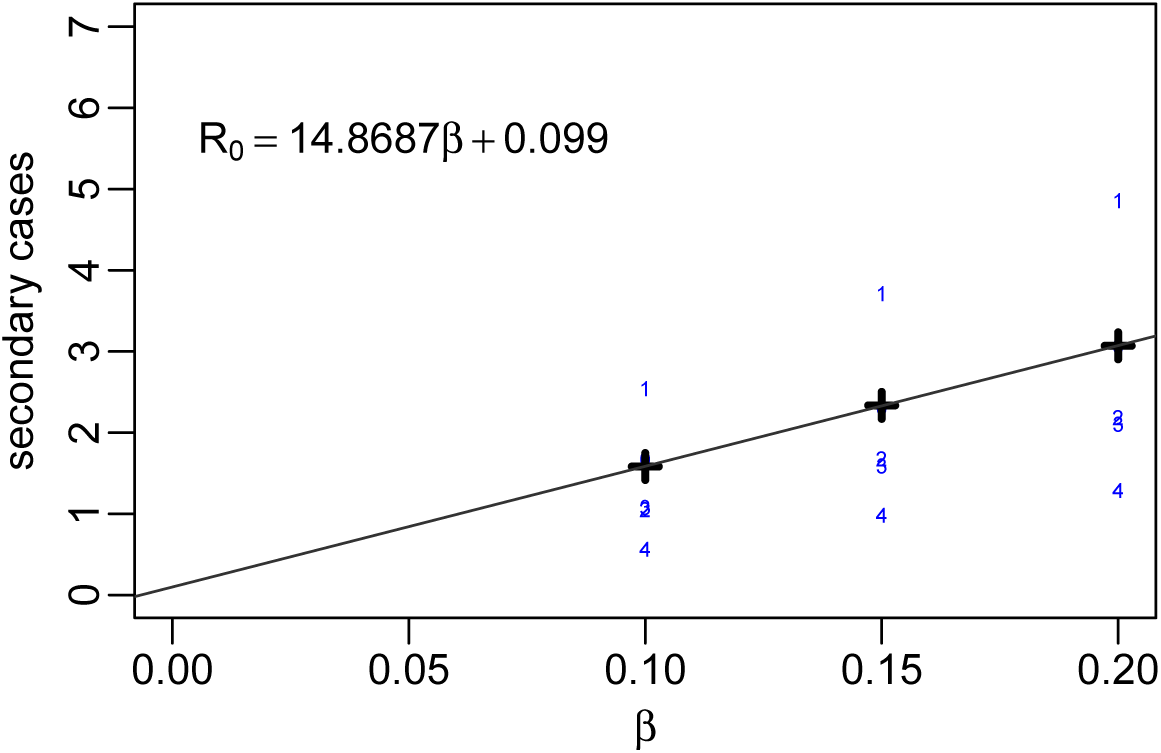
*β* vs R_0_ in the model. The numbers represent the age bin of the index case (0..4).

Things that affect R_0_ (and would require model recalibration if changed):

- virus’s incubation period
- symptomatic fraction
- infectiousness over time
- tendency of people to stay home when sick
- family structure/age structure of the population
- proportion of the population that goes to school or a workplace

## Data Availability

The source code for the model and scripts to generate the main figures are available at: https://github.com/dlchao/corvid

## Acknowledgments

DLC, APO, and MF would like to thank Bill and Melinda Gates for their active support of the Institute for Disease Modeling and their sponsorship through the Global Good Fund. We would also like to thank Edward Wenger for fruitful discussions and thoughtful reviews of early drafts and George Kour for the LaTeX template that we used to format this manuscript.

## A Supporting Material

### A.1 Characteristics of a simulated unmitigated outbreak

We ran simulations in a synthetic population that represents a large American city with a population of over 560,000. In an unmitigated outbreak starting with five infected people, 81% of the population became infected when R_0_=2.6 (Figure 10). The doubling time was initially 5.4 days before slowing 8.4 days, and the average generation time was 8.4 days, median of 8.0 days (Figure 11). The maximum infectious period allowed by the model is 21 days.

**Figure 10:**
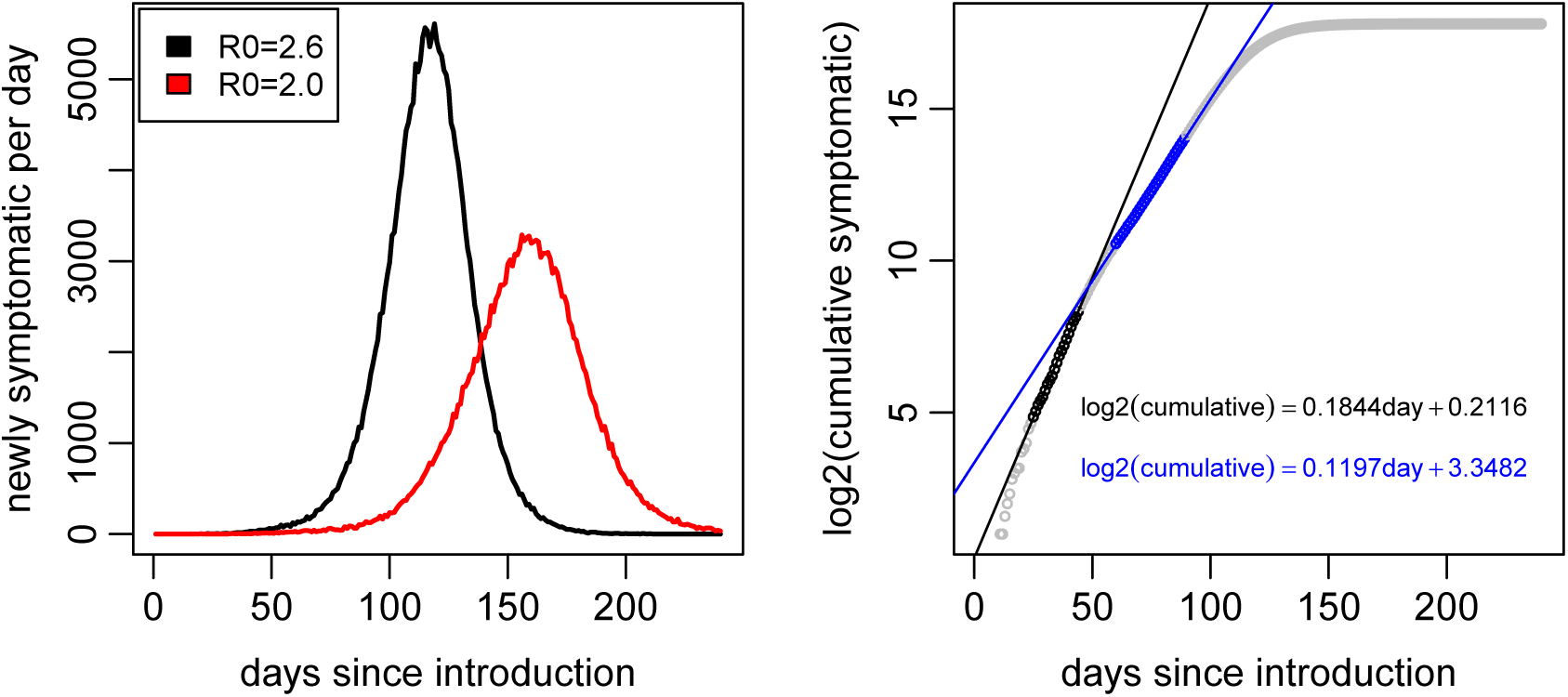
Symptomatic people over time in a metropolitan area in the model. On the left, the number of people who became symptomatic each day for R_0_=2.6 and R_0_=2.0. On the right, the log base 2 of cumulative cases plotted with two linear regression fits to estimate doubling time for R_0_=2.6.

**Figure 11:**
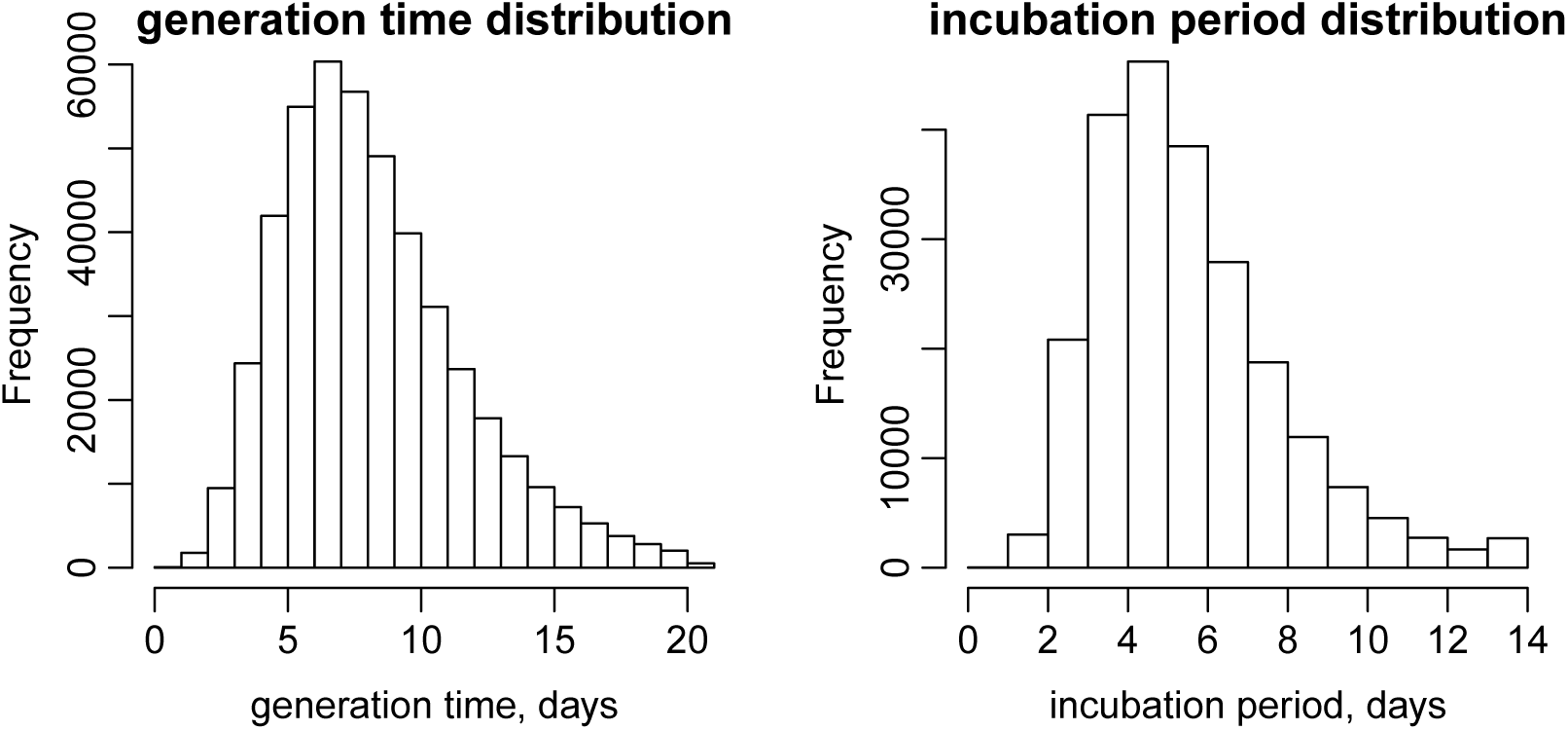
Generation time and incubation period distribution in the model. On the left, the distribution of generation times. On the right, the incubation period distribution. Results from one epidemic simulation with R_0_=2.6.

### A.2 Where do infections occur in the model?

In simulated epidemics, a large proportion of infections occurred within households (Table 1). School and workplaces contribute less but still comprise a significant share of transmission. The remaining infections come from the general community. For lower R_0_, schools may play a larger role in transmission. For high R_0_, the attack rate for all age groups is high, so the age distribution of cases flattens and adults (and settings with only adults) become more important.

In the model, if an individual becomes infected with SARS-CoV-2, there is a high probability that other family members will become infected. We can measure the household secondary attack rates by infecting a random individual in the population with a non-transmissible pathogen (using the method used to compute R_0_ as described in Section 4.2) and count the number of infections in that person’s household. For R_0_=2.6, if an index case is a child, the household infection attack rate is 51% for children and 28% for adults (Table 2). If an index case is an adult, the household infection attack rate is 22% for children and 32% for adults.

## Notes

### Competing Interest Statement

The authors have declared no competing interest.

